# Psilocybin-Assisted Group Psychotherapy + Mindfulness Based Stress Reduction (MBSR) for Frontline Healthcare Provider COVID-19 Related Depression and Burnout: A Randomized Clinical Trial

**DOI:** 10.1101/2024.12.31.24319806

**Authors:** Benjamin R. Lewis, John Hendrick, Kevin Byrne, Madeleine Odette, Chaorong Wu, Eric L. Garland

## Abstract

**Objective:** This clinical trial sought to evaluate the safety and preliminary efficacy of psilocybin and MBSR for frontline healthcare providers with symptoms of depression and burnout related to the COVID-19 pandemic.

**Methods:** This was a randomized controlled trial that enrolled physicians and nurses with frontline clinical work during the COVID-19 pandemic and symptoms of depression and burnout. Participants were randomized in a 1:1 ratio to either an 8-week MBSR curriculum alone or an 8-week MBSR curriculum plus group psilocybin-assisted psychotherapy (PAP) with 25mg psilocybin. Symptoms of depression and burnout were assessed at baseline, and 2-weeks and 6-months post intervention utilizing the Quick Inventory of Depressive Symptoms (QIDS-SR-16) and Maslach Burnout Inventory Human Services Survey for Medical Professionals (MBI-HSS-MP), respectively. Secondary outcome measures included the Demoralization Scale (DS-II) and the Watt’s Connectedness Scale (WCS). Adverse events and suicidality were assessed through 6-month follow-up.

**Results:** 25 participants were enrolled and randomized. There were 12 study-related AEs recorded that were Grade 1-2 and no serious AEs. There was larger decrease in QIDS score for the MBSR+PAP arm compared to MBSR-only from baseline to 2-weeks post-intervention and significant between-group differences favoring MBSR+PAP on subscales of the MBI-HSS-MP as well as the DS-II and WCS.

**Conclusions:** Group psilocybin-assisted therapy plus MBSR was associated with clinically significant improvement in depressive symptoms without serious adverse events and with greater reduction in symptoms than MBSR alone. Study findings suggest that integrating psilocybin with mindfulness training may represent a promising treatment for depression and burnout among physicians and nurses.

## INTRODUCTION

Depression and burnout among physicians and nurses have been recognized as worsening crises in the U.S. medical system. These issues have been exacerbated by the SARS-CoV-2 pandemic, where chronic, system-dependent stressors were coupled with dramatic increases in clinical demand, limited resources and resource rationing, assumption of increased personal risk, and increasing difficulties in balancing family life and professional responsibilities.(1–6) Burnout is a recognized psychological syndrome characterized by emotional exhaustion, depersonalization, and reduced personal accomplishment (7) and may lead to a sense of disconnection in the clinician-patient relationship. Mindfulness, a mental training practice involving present-moment, nonjudgmental awareness of thoughts and emotions, may be a promising means of addressing depression and burnout among healthcare providers.

Mindfulness-Based Stress Reduction (MBSR) is a well-established, evidence-based mindfulness training program that has been shown to reduce symptoms of depression, anxiety, and burnout as well as other mental health conditions among patients(8) and healthcare providers.(9–11) Similarly, psychedelics such as psilocybin have demonstrated efficacy in treating depressive symptoms.(12–14) There is increasing scientific interest in the potential synergy between mindfulness training and psychedelics.(15–22) Mindfulness and psychedelics appear to activate overlapping brain circuits(23) and theorists suggest that psychedelic experiences may deepen or help cultivate mindfulness skills.(16) Moreover, administering psychedelic-assisted therapy within the context of mindfulness training may lead to more durable therapeutic effects.

There has been one published randomized controlled trial of individual format psilocybin-assisted therapy for symptoms of depression and burnout in frontline healthcare providers.(24) This study demonstrated a significant reduction in depressive symptoms for the psilocybin treatment group at the 28-day follow up time point and suggested this treatment modality may be an effective intervention for providers dealing with depressive symptoms in the post-pandemic milieu.

However, this prior study – as with most studies of psilocybin-assisted psychotherapy (PAP) to date – did not involve an active treatment control and also employed an individual PAP format with a 2:1 therapist-to-participant ratio. This delivery format significantly limits scalability and accessibility of this resource-intensive treatment and precludes possible therapeutic aspects of group-based interventions for conditions (like depression and burnout) characterized in part by a sense of isolation and lack of connection. To date, there have been three prior psilocybin trials employing variations on group format interventions.(25–27) There are compelling reasons to hypothesize that group-based psilocybin-assisted psychotherapy (PAP) may offer a uniquely effective way of augmenting the benefits of mindfulness interventions as well as improving symptoms of burnout.

Here we conducted the first randomized controlled trial (RCT) of MBSR vs. MBSR + PAP in a group format for frontline physicians and nurses experiencing burnout and depression related to the COVID-19 pandemic. This study design employing an evidence-based psychotherapeutic intervention with an active control condition responds directly to recent recommendations by Seybert et al. who present a call to the field to clearly specify and examine optimal psychotherapeutic adjuncts to psilocybin treatments.(28)

## METHODS

### Study Design, Setting, Participants

This parallel randomized controlled trial (NCT05557643) investigated the safety and preliminary efficacy of MBSR+PAP vs. MBSR for health care providers with a DSM-5 depressive disorder and symptoms of burnout as measured by the Maslach Burnout Inventory Human Services Survey for Medical Professionals (MBI-HSS-MP). Patients were recruited from December, 2022 to February, 2024. The University of Utah institutional review board approved the protocol. The study was registered on www.clinicaltrials.gov: ClinicalTrials.gov Identifier: NCT05557643, https://clinicaltrials.gov/study/NCT05557643?term=NCT05557643&rank=1. All study processes were conducted at the University of Utah Huntsman Mental Health Institute.

Eligible participants were physicians (MDs) or nurses (RNs) with at least one month of frontline COVID-19 patient contact, who met DSM-5 criteria for a depressive disorder (PHQ-9 score ≥10) and had MBI-HSS-MP scores of ≥27 on the Emotional Exhaustion subscale and high scores on either the Depersonalization (≥13) or Personal Accomplishment subscales (≤ 21). Exclusion criteria included history of psychosis or mania, family history of first degree relative with a psychotic disorder, recent use of excluded psychiatric medications, active substance use disorder, and suicidal behavior. The Columbia Suicide Severity Rating Scale (C-SSRS) screening version was administered during screening to assess suicidality. Participants randomized to MBSR+PAP were required to taper existing antidepressant medications (n=2). After obtaining informed consent, coordinators collected demographic information.

### Masking and Randomization

Before randomization, participants completed a preference/credibility/expectancy assessment, metabolic panel, urine drug screen, and pregnancy test (if applicable). An investigator uninvolved in assessments or analysis generated treatment allocations to MBSR+PAP or MBSR with random assignment (1:1 ratio) in blocks of 3-5 per study arm per cohort. Treatment allocation was assigned to participants with sealed envelopes. Assessments were conducted as self-report measures by participants. To maintain blinding, the study key with allocations was inaccessible to the statistician until study completion. No placebo drug control was used and thus participants were not blinded to psychedelic treatment.

### Interventions

Participants enrolled were enrolled in a standard MBSR course, involving eight weekly, two-hour group sessions in which mindfulness meditation training (e.g., mindful breathing, body scan) and psychoeducation were provided.(29) For participants randomized to the MBSR+PAP arm, the psilocybin intervention began after four weeks of MBSR and included three group preparatory sessions over a one week period, a group dosing session (following established group PAP protocols)(25,30) and three group integration sessions over a two week period (**Figure 1)**. See **Supplement 2** for a full, detailed description of the group psilocybin intervention per recommendations by Seybert et al.(28) Each participant in the MBSR+PAP arm was paired with an individual therapist, and preparatory and integration sessions included a 30-minute one-on-one ‘break-out’ session with their assigned therapist. Group therapy followed a supportive-expressive model. Therapist engagement during the dosing session was supportive and nondirective. During psilocybin dosing, vital signs were monitored every 30 minutes for the first two hours, then hourly. Each participant completed AE assessments, the C-SSRS, and had a clinical evaluation to ensure safety prior to departure from the site. Participants in the MBSR-only arm attended an in-person all-day silent meditation retreat concurrent with the psilocybin dosing day. For the MBSR+PAP arm, three integration sessions were held on days 2, 6, and 13 post-dosing. The Group PAP Protocol can be found in the Supplementary Appendix.

**Figure 1.**
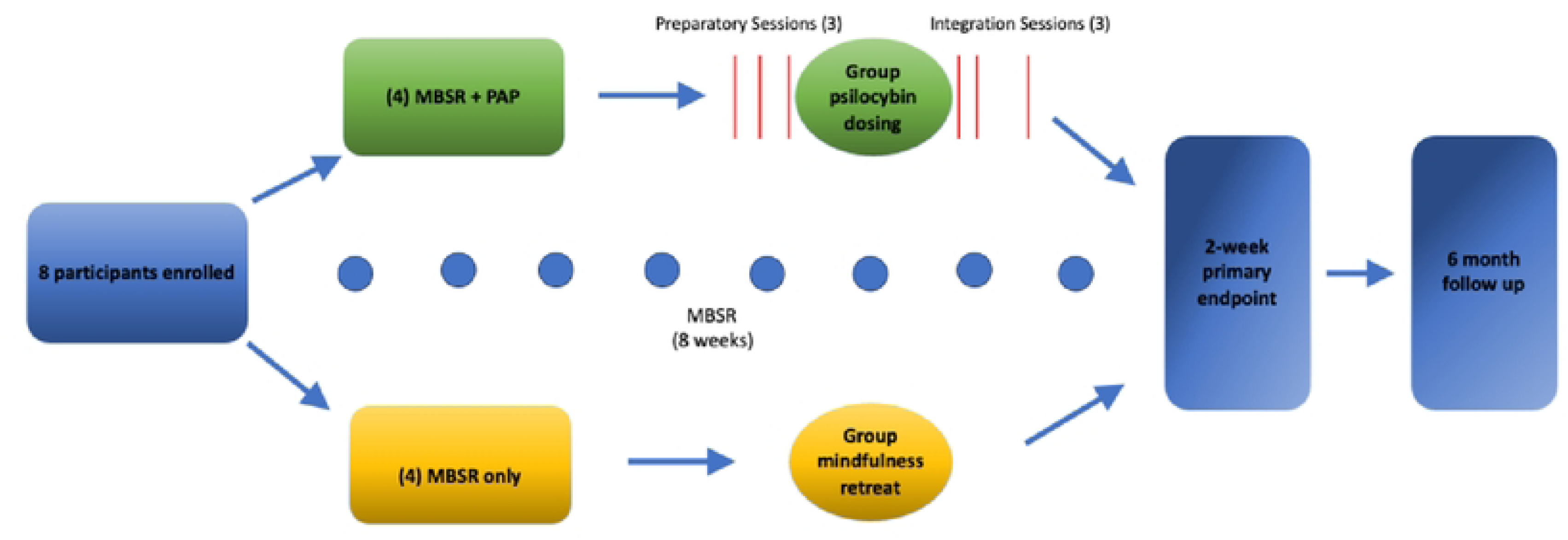
Study Flow Chart.

### Safety and Feasibility Outcomes

Feasibility was measured by recruitment, retention, and completion rates, with a target of at least 66% attendance at MBSR sessions for both study arms and 75% attendance at preparatory and integration sessions for the MBSR+PAP arm. AEs were evaluated at weeks 1, 3, 5, 6, 7, 8, 9, 11 and 6-months and categorized using Common Terminology Criteria for Adverse Events version 5 (CTCAE v.5). The C-SSRS was administered at screening, weeks 1, 6, 8, 9, 11, and 6-months.

### Clinical Outcomes

Outcome measures were collected at baseline, 2-weeks, and 6-months post-intervention. The primary clinical endpoint was reduction in Quick Inventory of Depressive Symptoms (QIDS-SR-16)(31) scores at 2-weeks post-intervention.

### Primary Outcome Measure Justification

The Quick Inventory of Depressive Symptomatology-Self Report (QIDS-SR-16) was chosen as the primary outcome measure for this study due to its robust psychometric properties, its wide acceptance in clinical and research settings, and its ease of administration. The QIDS-SR-16 is a 16-item brief, self-administered instrument designed to assess the severity of depressive symptoms along nine DSM-based symptom domains of major depressive disorder. It is translated into multiple languages (including Spanish, French, German, and Chinese). Each symptom domain is represented by 1-4 items, with scores ranging from 0-3 per domain and a total score of 0-27, with higher scores indicating more severe depressive symptoms. The QIDS-SR-16 has excellent reliability and validity across various populations with high internal consistency (Cronbach’s alpha typically exceeding 0.85) and correlates highly with other established clinician-administered depression measures such as the HAM-D.(31) There is a precedent for using this tool in previous studies with psilocybin-assisted therapy.(32) A 3.5 point reduction on the QIDS-SR-16 is considered a clinically significant reduction and total score can be interpreted using standard severity ranges (0-5 no depression, 6-10 mild depression, 11-15 moderate depression, 16-20 severe depression, 21-27 very severe depression). This established interpretability facilitates making meaningful conclusions regarding treatment effect. The self-reported format of this tool increases ease of administration, enhances autonomy, and minimizes observer bias. The QIDS-SR-16 is available free of charge for academic institutions with a Master User License Agreement through MAPI Trust (www.mapi-trust.org).

The key secondary clinical outcome was the MBI-HSS-MP, which includes 3 subscales: emotional exhaustion, depersonalization, and personal achievement.(33) Additional secondary outcomes included the Demoralization Scale (DS-II),(34) and the Watts Connectedness Scale, a self-report questionnaire measuring connectedness to self, others, and world that includes a General Connectedness Scale and subscales of Connectedness to Self (CTS), Connectedness to Others (CTO), and Connectedness to World (CTW).(35) Expectancy was assessed using the Credibility/Expectancy Questionnaire(36) prior to randomization and after participants learned of their random assignment. Experiential measures including the Mystical Experience Questionnaire (MEQ-30)(37), Challenging Experience Questionnaire (CEQ)(38), and NADA-state(39) were administered at the end of either the psilocybin dosing day or the MBSR meditation retreat to all participants.

### Statistical Analysis

A meta-analysis examining the effects of psilocybin on depressive symptoms reported an effect size of Cohen’s d=1.29. (40) Anticipating an 18% drop out rate (consistent with prior PAP studies), with an effect size of 1.0 and alpha=0.05, a sample size of 24 patients would provide 80% power to detect between-groups differences in the primary efficacy outcome.

The intent-to-treat analysis on all efficacy outcomes included all randomized participants (N=25). The per-protocol analysis (N=20) included participants in the MBSR arm who completed two-thirds of the MBSR sessions and, for those in the MBSR+PAR arm, psilocybin dosing and two-thirds of the preparatory and integration sessions. Analyses were conducted with linear mixed models (LMMs) with maximum likelihood estimation. Models specified random intercepts. Time point was treated as a categorical variable and the interaction between treatment arm and time point was included to evaluate between-group changes in outcomes over time. The effect of post-randomization expectancy on change in QIDS-SR-16 score from baseline to the 2-week endpoint was assessed using correlation analyses on both study arms.

The correlation between experiential scales (MEQ-30, CEQ, and NADA-state) were evaluated using linear regression and summarized with Pearson coefficients.

Statistical analyses were carried out using R 4.4.0 (R Core Team, 2024, with a significance level set at α = 0.05.

## RESULTS

### Participants

We assessed 420 patients for eligibility (**Figure 2**), and enrolled 25. Mean (SD) participant age was 40.4 (SD 8.4) in the MBSR-only arm and 47.4 (SD 10.9) in the MBSR+PAP arm. 72% of participants were women and the majority (96%) were white. Of the enrolled sample, 10 were MDs and 15 were RNs. The mean QIDS-SR-16 score at baseline was 12.3 (SD 3.9), indicating moderate depression(31) with no significant difference between study arms. The majority (88%) of participants had a lifetime history of antidepressant use. One participant dropped out of the MBSR+PAP arm and two participants dropped out of the MBSR-only arm (**Figure 2**) due to inability to adhere to the time commitments. No participants withdrew due to an AE. There was no significant between-groups difference in preference for study arm assignment. Baseline characteristics of the intent-to-treat (ITT) sample are provided in **Table 1**.

**Figure 2.**
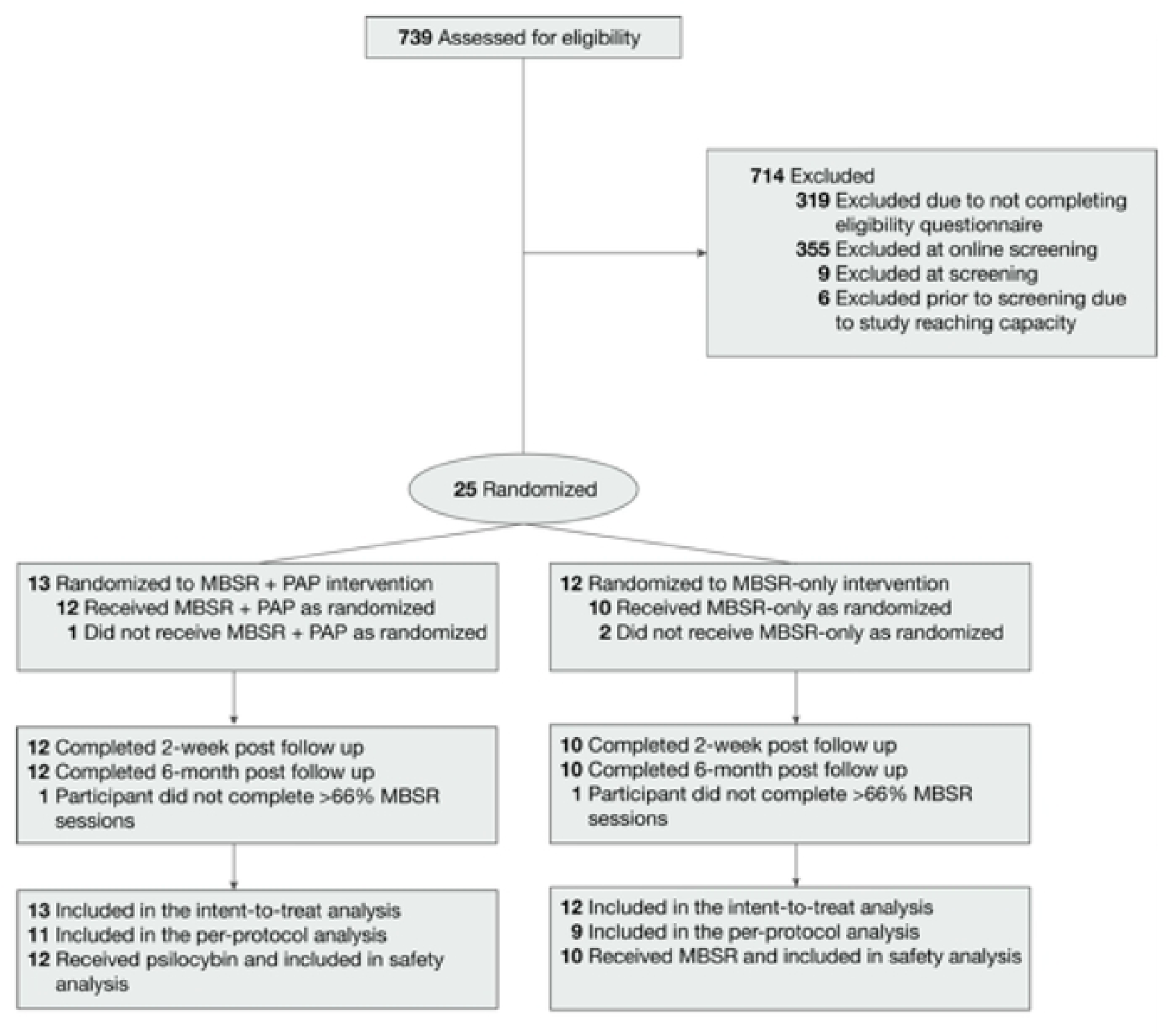
Enrollment, Randomization, and Follow-up of Participants.

**Table 1.** Demographic and Clinical Characteristics of Participants at Baseline. (1) As determined by chart review and screening visit MD assessment. (2) Two participants randomized to MBSR+PAP arm required tapering of antidepressants. Participants randomized to MBSR-only were not required to taper existing antidepressant treatments. Abbreviations: QIDS-SR-16 = Quick Inventory of Depressive Symptomatology – Self-Report (16-item); MBI (EE), Emotional Exhaustion Subscale of MBI. MBI (DP), Depersonalization Subscale of MBI. MBI (PA), Personal Accomplishment Subscale of MBI.

| Category | Characteristic | MBSR only (n=12) | MBSR + PAP (n=13) |
| --- | --- | --- | --- |
| Demographics | Age– mean (range) | 40 (32-49) | 47 (36-58) |
|  | Female sex– no. (%) | 6 (50%) | 12 (92%) |
|  | White race– no. (%) | 11 (92%) | 13 (100%) |
| Specialty | MD no. (%) | 5 (42%) | 5 (38%) |
|  | RN no. (%) | 7 (58%) | 8 (62%) |
|  | Emergency Medicine | 2 | 4 |
|  | Oncology | 2 | 1 |
|  | Palliative Care | 0 | 1 |
|  | Psychiatry | 1 | 3 |
|  | Anesthesiology | 3 | 0 |
|  | Critical Care | 1 | 1 |
|  | Surgery | 1 | 0 |
|  | Urology | 1 | 0 |
|  | Pediatrics | 0 | 1 |
|  | Neurology | 1 | 0 |
|  | Internal Medicine | 1 | 0 |
|  | Primary Care | 0 | 1 |
| Psychiatric Diagnoses (1) | Major depressive disorder– no. (%) | 11 (92%) | 13 (100%) |
|  | Adjustment disorder, with depressed mood– no. (%) | 1 (8%) | 0 |
|  | Adjustment disorder, with mixed anxiety and depressed mood– no. (%) | 0 | 0 |
| Treatment and Substance Use History | Prior Psychotropic Medication– no. (%) (2) | 10 (83%) | 12 (92%) |
|  | Prior Psychedelic Use– no. (%) | 6 (50%) | 6 (46%) |
|  | Current Cannabis Use– no. (%) | 4 (33%) | 5 (38%) |
|  | Current Alcohol Use– no. (%) | 8 (67%) | 10 (77%) |
| Baseline Depression and Burnout Scores | QIDS-SR-16 – (SD) | 12.5 (2.9) | 12.1 (4.7) |
|  | MBI(EE), MBI(DP), MBI(PA) -(SD) | 42.8(7.4), 16(7.1), 31.7(7.8) | 42.2(8.6), 17.8(6.2), 28.4(8.8) |

### Safety and Feasibility

All study-related AEs were Grade 1 or 2 per CTCAE v.5.0 categorization, and 12 related AEs were reported (**Table 2**). There were no incidences of emergent suicidality or self-injurious behaviors in either study arm. There were no clinically significant changes in vital signs during psilocybin dosing that required emergent PRN antihypertensive use. The three reported instances of nausea during the psilocybin session were self-limited and did not require anti-emetic administration. There were no administrations of pro re nata (PRN) lorazepam for acute anxiety.

**Table 2.** Study-Related Adverse Events.

|  | **MBSR (n=12)**<br>**No. (%)** | **PAP (n=13)**<br>**No. (%)** |
| --- | --- | --- |
| At least one AE | 4 (33) | 13 (100) |
| At least one related AE | 2 (17) | 6 (46) |
| At least one serious AE | 0 (0) | 0 (0) |
| **AE severity** |  |  |
| Mild | 6 | 9 |
| Moderate | 7 | 39 |
| Severe | 0 | 0 |
| **Study-related AE severity** |  |  |
| Mild | 1 | 11 |
| Moderate | 1 | 3 |
| Severe | 0 | 0 |
| **Study-related AEs** |  |  |
| Headache | 0 | 4 |
| Anxiety | 2 | 3 |
| Nausea | 0 | 2 |
| Hot flashes | 0 | 1 |
| Marital conflict | 0 | 1 |
| Dizziness | 0 | 1 |
| Rhinorrea/lacrimation | 0 | 1 |
| Malaise (related to stopping SSRI) | 0 | 1 |
| **Drug-related AEs** |  |  |
| Headache | NA | 3 |
| Nausea | NA | 2 |
| Hot flashes | NA | 1 |
| Anxiety | NA | 1 |
| Dizziness | NA | 1 |
| Rhinorrea/lacrimation | NA | 1 |

In the MBSR+PAP arm there was 100% attendance of the three preparatory sessions, 100% attendance of the dosing session, and 97.2% attendance of the three integration sessions. Attendance of scheduled MBSR sessions was 80.9% across both arms; two participants did not attend at least two-thirds of scheduled MBSR sessions and were not included in the per-protocol analysis (**eTable 9**, **Supplement 1**).

### Preliminary Efficacy

The MBSR+PAP arm evidenced significantly greater reduction in QIDS-SR-16 scores than the MBSR-only arm from baseline to the primary 2-weeks post-intervention endpoint between-groups effect=4.6, 95% CI=1.51-7.70, p=0.004, d=1.04). A 3.5 point decrease is considered a clinically significant reduction on the QIDS-SR-16(31); MBSR+PAP reduced QIDS-SR-16 scores by 7.26 points (SE=1.04) compared with a 2.66 point (SE=1.14) reduction in the MBSR-only condition at the 2-week endpoint. Mean QIDS-SR-16 score at the 2-week endpoint was 4.75 (SD=2.4) for the MBSR+PAP condition: a score of 5 or lower on this scale is considered evidence of no depression (**Table 3**, **Figure 3**).

**Figure 3:**
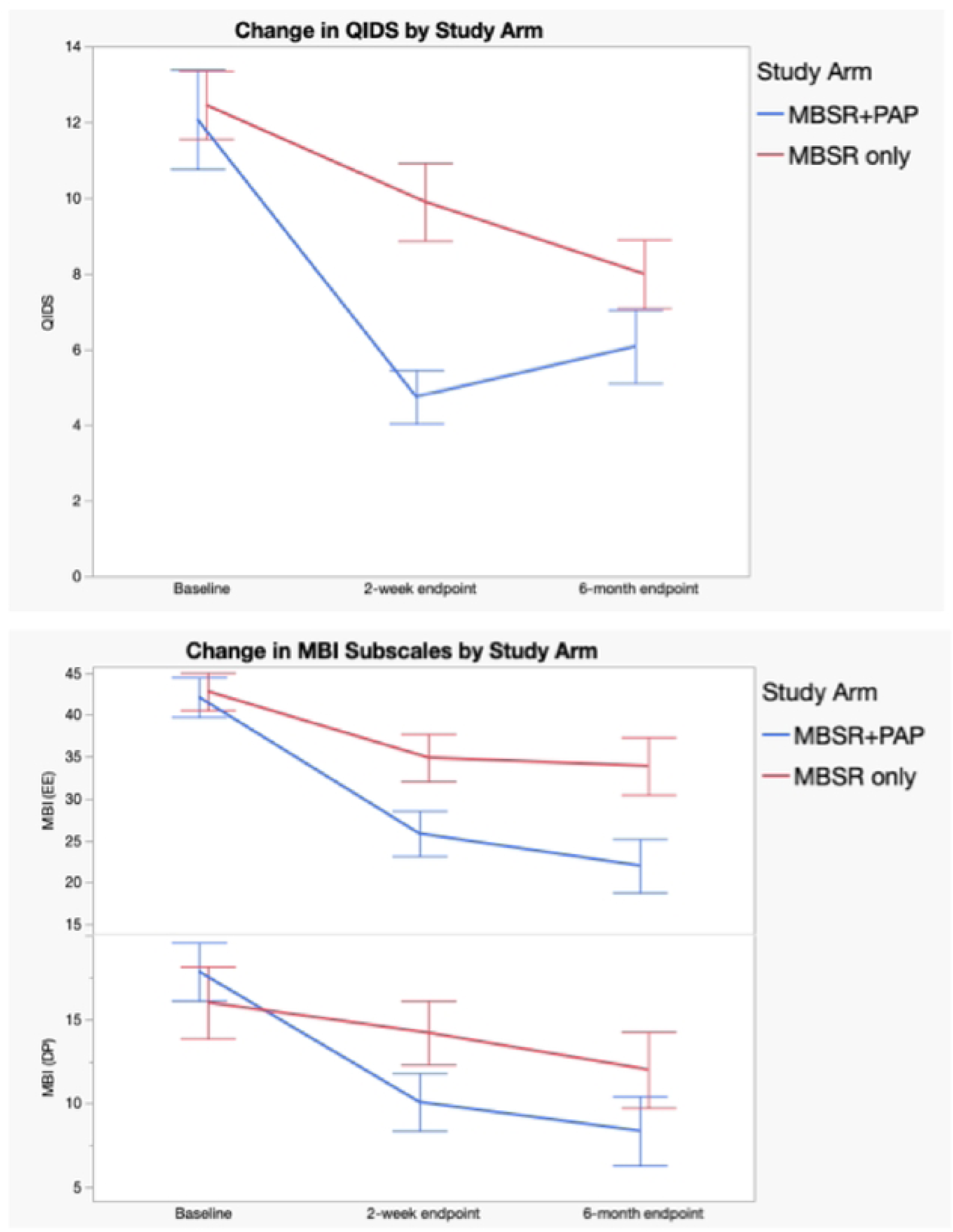
Efficacy Outcomes. Change in Quick Inventory of Depressive Symptoms (QIDS-SR-16) Score and Change in Maslach Burnout Inventory (MBI-HSS-MP) Emotional Exhaustion (EE) and Depersonalization (DP) Subscale Scores by Treatment Group (Intention-to-Treat Analysis). Total scores on the Quick Inventory of Depressive Symptoms range from 0-27 with higher scores indicating greater severity of depression. Bars represent standard errors.

**Table 3.**
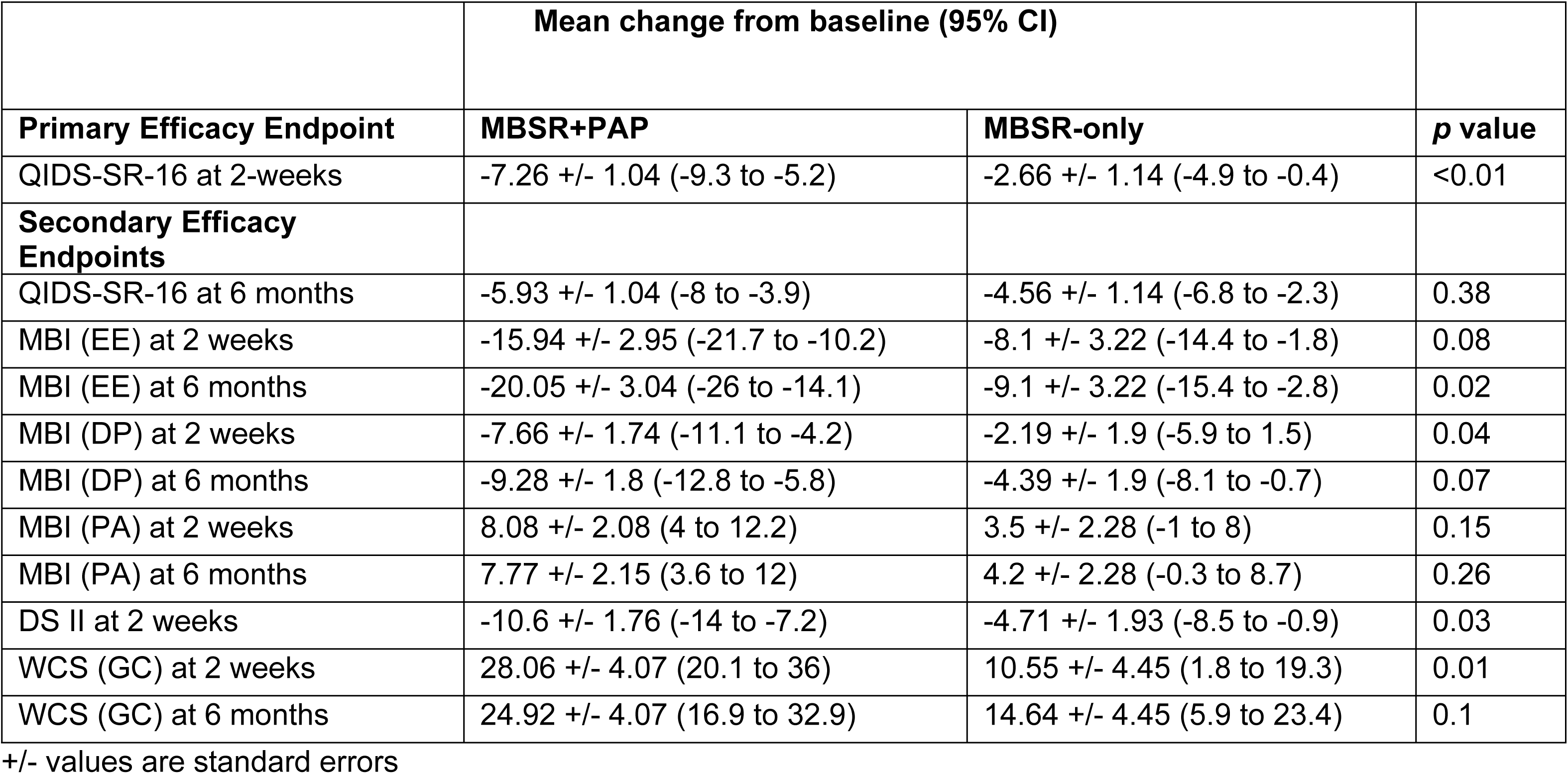
Primary and Secondary Efficacy End Points (Intention-to-Treat Population). Abbreviations: QIDS, Quick Inventory of Depressive Symptoms-16 item Self Report. MBI, Maslach Burnout Inventory Human Services Survey for Medical Professionals. MBI (EE), Emotional Exhaustion Subscale of MBI. MBI (DP), Depersonalization Subscale of MBI. MBI (PA), Personal Accomplishment Subscale of MBI. DS-II, Demoralization II Scale. NADA-trait, Nondual Awareness Dimensional Assessment, trait. WCS (GC), Watt’s Connectedness Scale General Connectedness measure. *p*-values represent Group x Time interactions from mixed model analyses.

Regarding secondary outcomes (**Table 3),** the MBSR+PAP arm demonstrated significantly greater reductions in the depersonalization subscale of the MBI-HSS-MP from baseline to 2-weeks post-intervention (between-groups effect=5.47, 95% CI= 0.3-10.6, p=0.038), and in emotional exhaustion from baseline to 6-months post-intervention (between-groups effect =10.9, 95% CI= 2.1-19.8, p=0.016). There were no significant between group differences on the personal accomplishment subscale due to ceiling effects from high scores at baseline. (**Figure 3**). MBSR+PAP outperformed MBSR-only in reducing demoralization from baseline to the 2-week endpoint (**Figure 4**). There were significant between-group differences on the measure of General Connectedness (a global measure of the 3 WCS subscales) favoring MBSR+PAP from baseline to 2-weeks (between-groups effect -17.5, 95% CI =-29.6 to -5.5, p=0.005) as well as significant between-group differences on CTS and CTO subscales (**Figures 5 and 6**). There were significant treatment x time interactions across all three time points (baseline, 2-week endpoint, 6-month endpoint) for the QIDS-SR-16, MBI(EE), WCS (General Connectedness) as well as WCS (CTS) and WCS (CTO) subscales (**eTable5 Supplement 1, Figure 6**).

**Figure 4.**
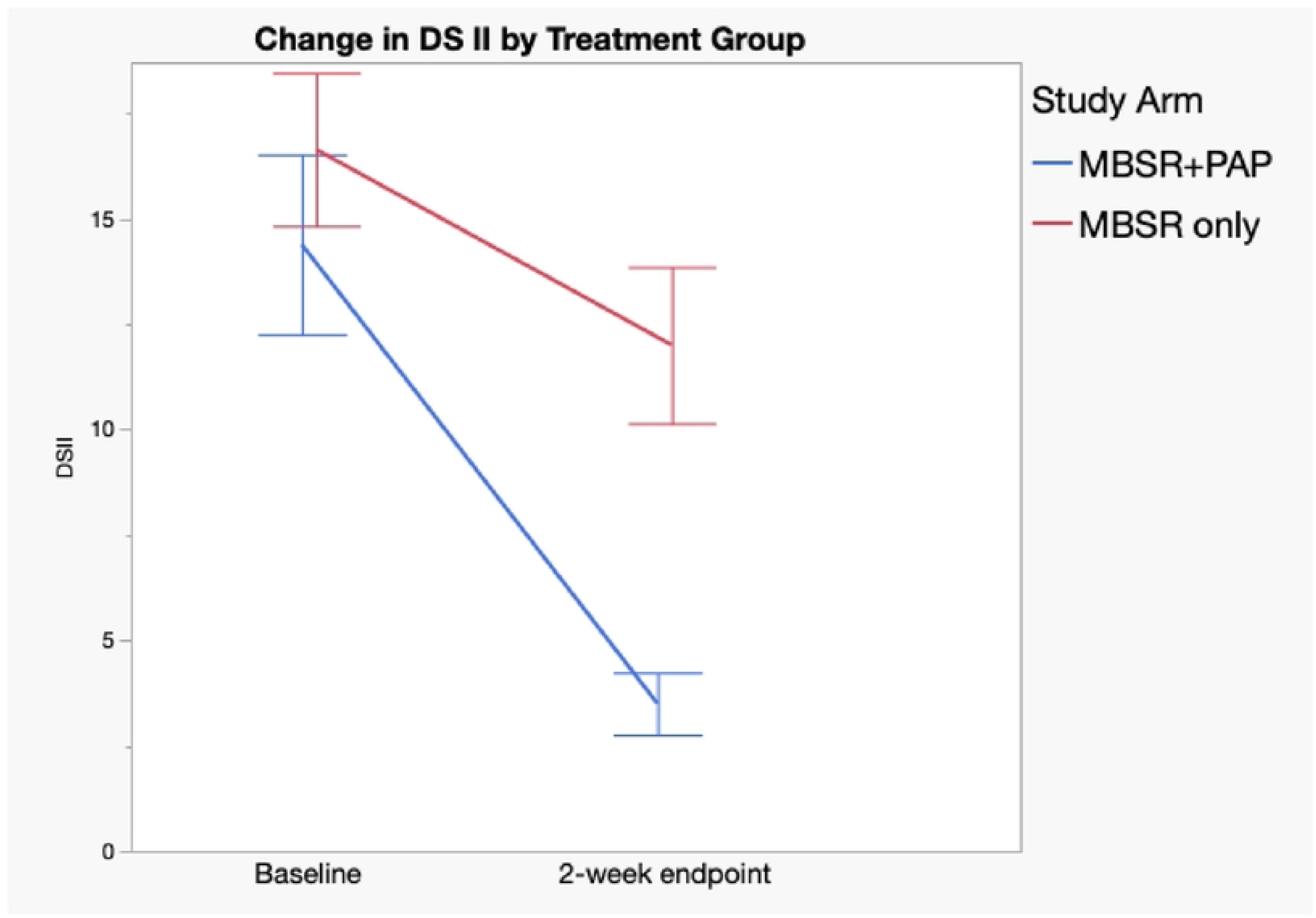
Change in Demoralization Scale (DSII) by Treatment Group. Assessed only at baseline and 2-week Endpoint. Error bars = standard error.

**Figure 5:**
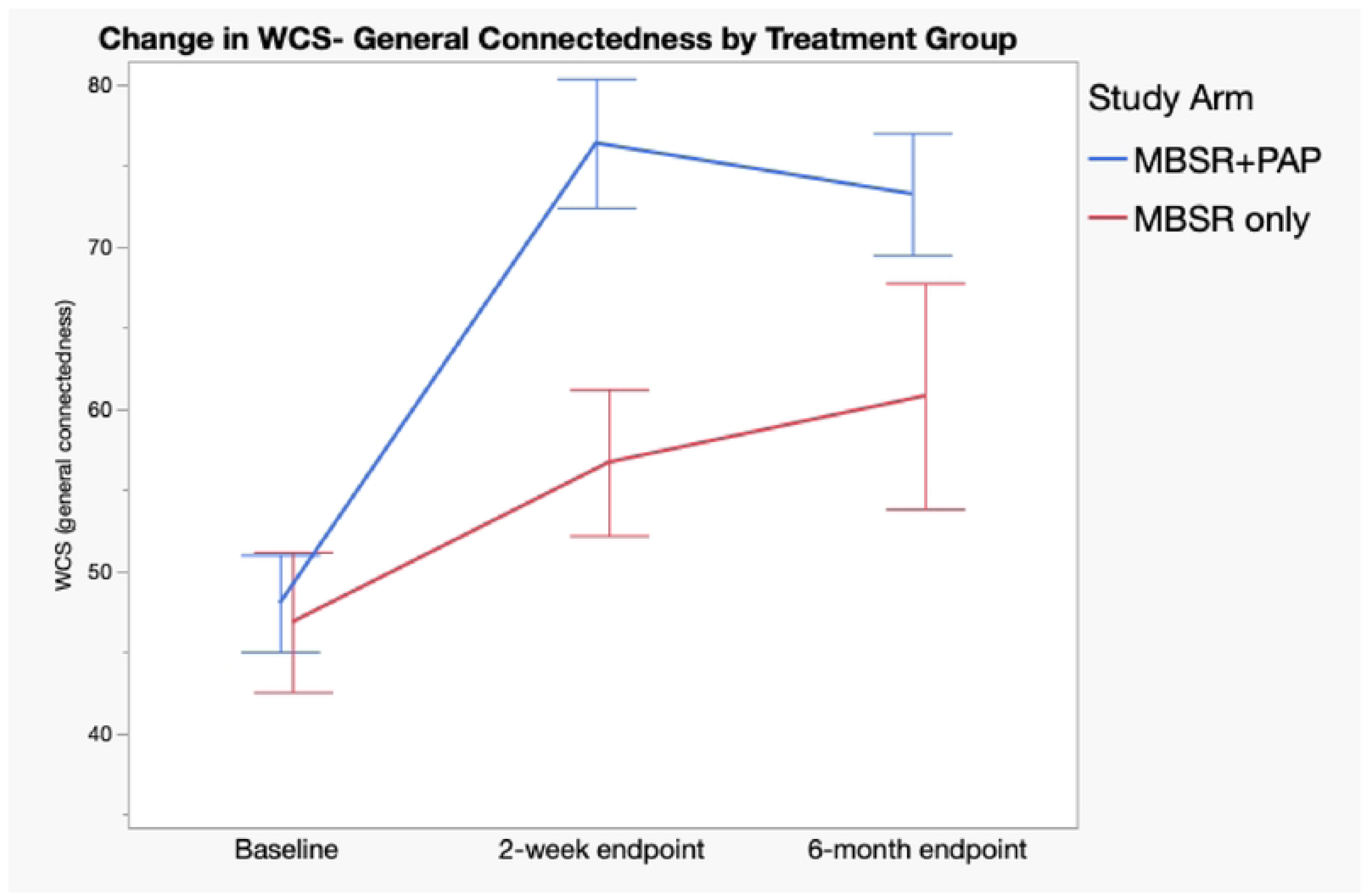
Change in Watt’s Connectedness Scale (WCS), General Connectedness by Treatment Group. General Connectedness measure= sum of subscales Connectedness to Self (CTS), Connectedness to Others (CTO), Connectedness to World (CTW). Error bars = standard error.

**Figure 6.**
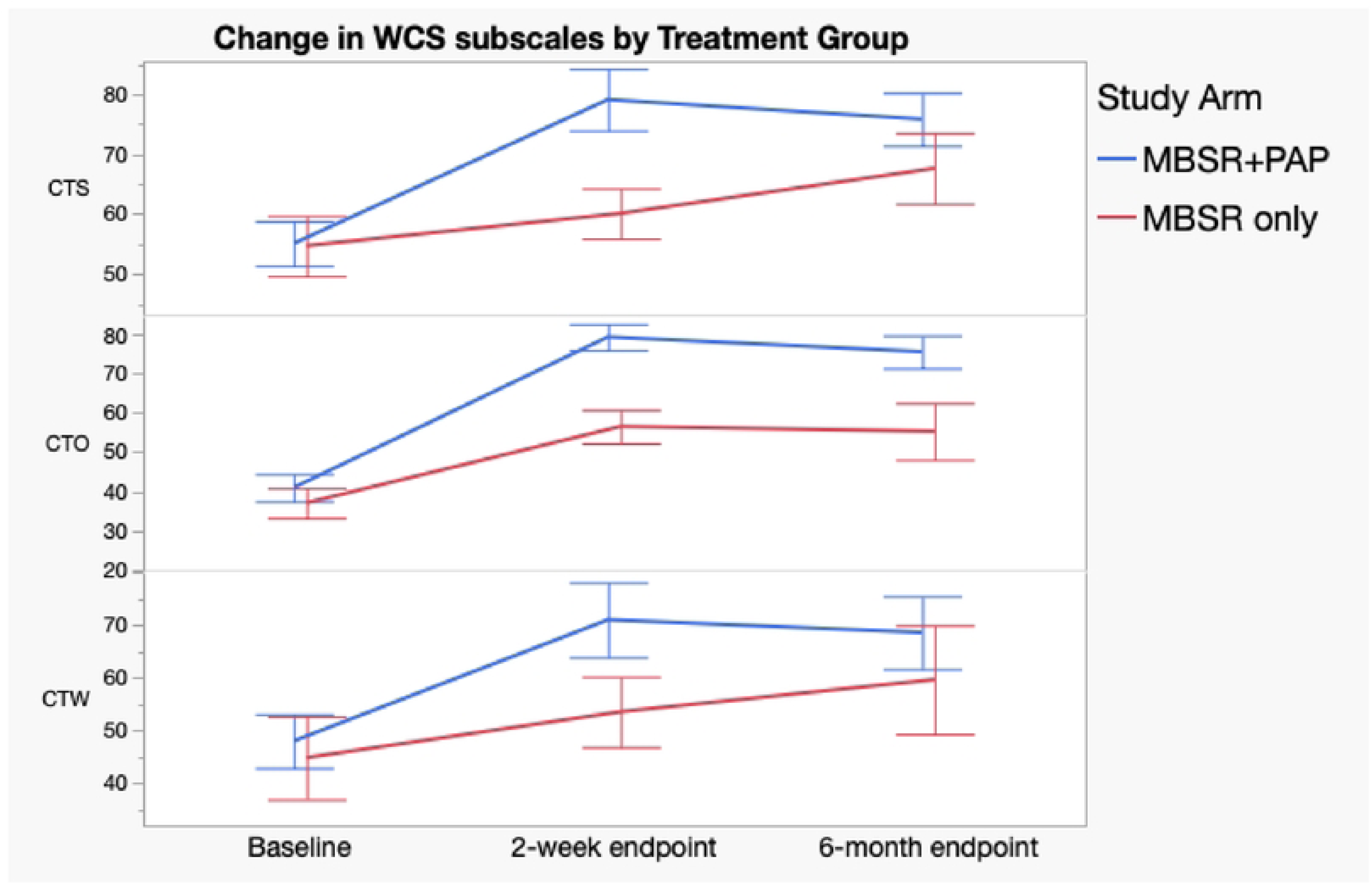
Change in Watt’s Connectedness Scale, Subscales by Treatment Group. CTS =Connection to Self, CTO= Connection to Others, CTW= Connection to World. Error bars = standard error.

There was a statistically significant difference (p=0.0003) in post-randomization expectancy for the MBSR+PAP arm (65.4, SD=14.7) vs. MBSR-only (37.6, SD=17.9). Expectancy was strongly associated with QIDS-SR-16 depression symptom score reduction in the MBSR-only arm (r=-0.70, p=0.022) but not in the MBSR+PAP arm (r=0.04, p=0.90). (**eTable 7**, **Supplement 1**).

There were no significant correlations between expectancy and change in MBI subscales at the 2-week endpoint.

Experiential questionnaires (MEQ-30, CEQ, and NADA-state) were administered to both study arms at either the end of the psilocybin dosing day (hour 7-8) or end of the MBSR retreat day. Mean MEQ score was 112.5 (SD 26.1) for the MBSR+PAP arm and 24.5 (SD 34.5) for the MBSR-only arm. Mean total CEQ score (scale 0-5) was 1.46 (SD 1.15) for the MBSR+PAP arm and 0.39 (SD 0.39) for the MBSR-only arm. Mean NADA-state score was 22 (SD 7.63) for the MBSR+PAP arm and 8.3 (SD 9.25) for the MBSR-only arm. 8/12 participants in the MBSR+PAP arm had a ‘complete mystical experience’ on the MEQ-30 (≥ 60% on all subscales) compared to 0/10 participants who completed the MEQ-30 in the MBSR-only arm.

There was a large overall correlation between magnitude of score on the MEQ-30 and NADA-state and change in QIDS-SR-16 scores from baseline to the 2-week endpoint (r=-0.62, p=0.0019 for MEQ-30, r=-0.65, p=0.0018 for NADA-state) however there were no significant between group differences. Similarly, we found significant correlations between MEQ-30 scores and change in MBI(EE) (r=-0.47, p= 0.0286), MBI(DP) (r=-0.044, p= 0.0421), and WCS (r=0.616, p= 0.0023) scores from baseline to the 2-week endpoint (**eTable 8**, **Supplement 1**). There were no significant correlations found between CEQ outcomes and change in primary or secondary outcome measures from baseline to the 2-week endpoint.

## DISCUSSION

This randomized clinical trial demonstrated the safety and preliminary efficacy of 25 mg psilocybin administered in group format in conjunction with an 8-week MBSR curriculum for physicians and nurses experiencing depression and burnout related to COVID-19. There were no serious treatment-emergent AEs through the course of the trial and no emergent suicidality or self-injurious behaviors. Meanwhile, MBSR+PAP was associated with clinically and statistically significant decreases in depressive symptoms and burnout, reduced demoralization, and significant increases in the sense of connectedness.

The observed effect size of MBSR+PAP on depression scores is consistent with previously reported psilocybin effect sizes on depressive symptoms.(40) We observed a large antidepressant effect at the 2-week endpoint. This finding contributes to the growing evidence base that psilocybin is a rapid-acting treatment for depression (13,14) and adds new evidence for efficacy in the unique population of MDs and RNs. This result is consistent with recently reported outcomes by Back et al who have looked at psilocybin-assisted therapy alone in individual format for a similar population.(24) MBSR+PAP also appeared to reduce emotional exhaustion and depersonalization, two key facets of burnout: this suggests the possibility of additional therapeutic aspects of the incorporation of mindfulness training as well as a group format design for this set of symptoms given the lack of statistically significant effects in burnout symptoms in the recent Back et al study.(24) Though current understanding conceptualized burnout and depression as different but overlapping conditions, the two conditions are thought to have reciprocal relationship.(7) While interventions such as MBSR and psilocybin may specifically target individual resilience and psychological flexibility(41,42), they do not necessarily address other possible factors mediating burnout such as adjusting workload demands, time management skills, and conflict resolution skills. It may be the case that these respective interventions address certain internal causal factors but not relevant systemic factors. Notably, the antidepressant effects of PAP were strongest at the 2-week endpoint. By the 6-month endpoint depression scores for participants in MBSR-only approached those of participants in MBSR+PAP, suggesting that psilocybin may accelerate the therapeutic benefits of mindfulness.(43)

Notably, we also observed significant effects of MBSR+PAP on participants’ sense of connectedness to self and others. Research on depression and burnout has highlighted the profound effects that social connection and social relationships have on the development as well as the resolution of these syndromes. Indeed, burnout undermines the clinician-patient relationship by reducing empathy and compassion.(44) The utilization of a group model for the intervention intentionally recognizes these social factors. Prior studies of group format psilocybin-assisted therapy—while small and preliminary—have suggested synergistic effects between group connectedness and therapeutic outcomes.(30) Group models also dramatically increase the scale on which these resource intensive treatments could be delivered.(45)

While there was clear preference for randomization to the MBSR+PAP arm, and higher-rated expectancy in the MBSR+PAP arm than MBSR-only, there was no indication that expectancy effects post-randomization were significantly associated with improvement with the psilocybin condition. Rather, we found a significant association in the MBSR-only condition. This is worth noting, given recent concerns regarding the effects of expectancy, functional unblinding, and confirmation bias in trials of psychedelic-assisted therapies.(46) This also aligns with a recent analysis of expectancy effects in a phase-2 RCT comparing escitalopram to psilocybin for major depressive disorder. (47) These results support prior suggestions(47) that expectancy bias may play a less significant role in the therapeutic effects of psilocybin-assisted therapy than previously suspected.

The MEQ-30, along with the NADA and CEQ were administered to all participants after either at the end of the psilocybin dosing day or MBSR retreat depending on randomization. Magnitude of score on the MEQ-30 was strongly correlated with improved outcomes at 2-weeks on the QIDS-SR-16, MBI(EE), MBI(DP), and WCS scales across the whole study sample. While there were notable between-group differences in mean scores on experiential scales notably the magnitude of mystical experience correlated with outcomes independent of psilocybin and there was no clear effect of study arm on this relationship. Previous studies of psilocybin-assisted therapy have demonstrated a relationship between magnitude of mystical experience on the MEQ-30 and clinical outcomes. (48) Demonstrating this effect independent of psilocybin administration supports the possibility that self-transcendent states, occasioned by flexible means including mindfulness, have salutary effects.(43)

This clinical trial had several important limitations. The small sample size limited statistical power and generalizability. The homogeneity of our sample, consisting predominantly of white female participants, further restricts the generalizability of our findings to more diverse populations. Our study design, while employing an active behavioral treatment (MBSR) as a control condition, was not blinded, and this may have contributed to the different effects across study arms. The interventions differed between arms, with the PAP group participating in a psilocybin dosing day, while the MBSR-only group attended a silent meditation retreat. This design ensured that both groups received a form of intensive experience, although the nature of these experiences were not equivalent. The study was also limited in that we did not exclude participants based on prior psychedelic experience (six participants in each arm had previously used psychedelics). The effects of PAP may differ between psychedelic-naïve individuals and those with prior experience; the impact of prior psychedelic use on treatment outcomes remains unclear. To more effectively characterize the contributions of PAP vs. MBSR, we recommend that future studies consider a double-blind RCT design with an active placebo or a full factorial study design to disentangle the independent and interactive effects of psilocybin and mindfulness training.

In conclusion, combining MBSR with psilocybin appears to be a safe, feasible, and potentially efficacious approach to addressing depression and burnout among frontline healthcare workers. Larger, more diverse, multi-site studies with placebo controls are needed to further evaluate the efficacy of integrating psychedelics and mindfulness interventions for clinician wellbeing.

## Data Availability

All data will be made available via a data sharing agreement after acceptance. Supporting information files include relevant data and analyses and these have been submitted. Raw data from the study is stored in REDCap and can be made available following acceptance.

## Study Drug Supply

Usona Institute

## Disclosures

Benjamin Lewis MD is an investigator on 2 industry sponsored trials that are being conducted at the Huntsman Mental Health Institute:

1. *Title: A Multicenter, Randomized, Double-Blind, Parallel-Group Dose-Controlled Study Evaluating the Safety and Efficacy of RE104 for Injection in the Treatment of Patients with Postpartum Depression (PPD)* Major Goals: Evaluate the efficacy of RE104, a novel short acting psychedelic agent, for women with postpartum depression. Status of Support: Active Project Number: NCT06342310 Name of PD/PI: Benjamin R. Lewis MD Role: Principal Investigator Source of Support: Reunion Neuroscience Primary Place of Performance: University of Utah, multisite trial Project/Proposal Start and End Date: 06/2024 – 09/2025 Total Award Amount (including Indirect Costs): per enrollment Person Months (Calendar/Academic/Summer) per budget period. Year (YYYY) Person Months 1. 2024 - 25 1.2 2. 2025 - 26 1.2
2. *Title: A phase III, multicenter, randomized, double blind, controlled study to investigate the efficacy, safety, and tolerability of two initial administrations of COMP360 in participants with treatment resistant depression*

Major Goals: Evaluate the efficacy of psilocybin administration with psychological support for individuals with treatment resistant depression.

Status of Support: Active Project Number: NCT05711940

Name of PD/PI: Brian Mickey MD PhD Role: Co-Investigator

Source of Support: COMPASS Pathways

Primary Place of Performance: Huntsman Mental Health Institute, University of Utah, multisite trial

Project/Proposal Start and End Date: 06/2024 – 09/2026 Total Award Amount (including Indirect Costs): per enrollment

Person Months (Calendar/Academic/Summer) per budget period.

1. 2024 - 25 1.8

2. 2025 - 26 1.8

Eric Garland, PhD is the Director of UCSD ONEMIND (Optimized Neuroscience-Enhanced Mindfulness Intervention Design). UCSD ONEMIND provides Mindfulness-Oriented Recovery Enhancement (MORE), mindfulness-based therapy, and cognitive behavioral therapy in the context of research trials for no cost to research participants; however, Dr. Garland has received honoraria and payment for delivering seminars, lectures, and teaching engagements (related to training clinicians in MORE), including those sponsored by institutions of higher education, government agencies, academic teaching hospitals, and medical centers. Dr. Garland also receives royalties from the sale of books related to MORE. Dr. Garland has also been a consultant and licensor to BehaVR, LLC.

